# MASK MANDATES REDUCE COVID-19 MORTALITY: Analysis of 37 States and the District of Columbia, with a further analysis of the impact of demographic and medical factors on efficacy

**DOI:** 10.1101/2021.05.09.21256922

**Authors:** Michael J. Maloney

**Affiliations:** Proof School, Department of Mathematics, 973 Mission St., San Francisco, CA 941

**Keywords:** COVID-19, MORTALITY, MASK MANDATE, SOCIAL CAPITAL, PUBLIC HEALTH POLICY, GOVERNMENT POLICY

## Abstract

As the number of COVID-19 deaths in the US increased, various policies were enacted to slow the spread of the pandemic. While the situation has improved in recent months, determining how best to combat the current pandemic is still essential. Failure to do so invites both further resurgences of the current pandemic, and more pandemics in the years to come. As a result of the widespread failure to contain the spread of COVID-19, enough deaths have occurred that the impact of policy on mortality may be statistically evaluated. This paper uses Optimal Discriminant Analysis (ODA) to evaluate the hypothesized ability of limited mask mandates (MM) to reduce the daily number of COVID-19 deaths in the states analyzed. The mandates were found to reduce mortality in half the states analyzed and did not result in increased mortality in any states. A full range of cofactors were analyzed to determine which, if any, influenced the efficacy of the mandates in the states in which mandates had an effect. Institutional Health Subindex of the Social Capital Index, state health score, population density, portion of the population with nongroup health insurance, state GDP, and the rate of pregnancy related diabetes were all correlated with increased mandate efficacy. In contrast, incarceration rate, overcrowded housing, severely overcrowded housing, portion of the population with military provided insurance, portion of the population uninsured, the portion of the population unable to see a doctor due to cost, and the portion of the population who were American Indian/Native Alaskan were all correlated with reduced mandate efficacy.

## INTRODUCTION

At the time of writing the SARS-CoV-2 virus, the cause of COVID-19, has infected 32,356,034 individuals in the United States (9.7% of the population) and killed 576,238 Americans. While 149,462,265 Americans have been vaccinated, only an estimated 32.8% of the population is fully vaccinated ^1^. It is estimated that 69.6% of the US population will need to be immune to achieve herd immunity ^2^.

While immunity builds in the public, policies that reduce mortality remain essential, as does a deeper understanding of the cofactors that strengthen or weaken the impact of those policies.

Several studies have demonstrated the efficacy of Governmental Mask Mandates promoting the wearing of face masks in public ^3^, in reducing infections ^4, 5, 6, 7, 8, 9, 10, 11, 12, 13,14^, and reducing mortality ^15, 16^, associated with COVID-19.

Several medical comorbidities have been associated with increased COVID-19 mortality including cardiovascular disease, obesity, diabetes, and pulmonary disease ^17, 18, 19, 20, 21, 22^. Demographic factors such as age, rate of nursing home occupancy, rates of incarceration, access to health care, population density and overcrowded housing have been cited as risk factors with COVID-19 ^23,24^. Socioeconomic and clinical cofactors are reported by some to account for racial variations in mortality ^25, 26, 27^ whereas other research supports an impact of race as an independent variable ^28^. Finally, Social Capital Index has been shown to impact both the rate of infection and mortality of COVID-19 ^29, 30, 31, 32^, and fractious partisanship has been shown to worsen the impact of the pandemic ^33^. A total of 62 putative cofactors were analyzed to examine how they impact Mask Mandates’ effect on COVID-19 mortality (See **Table 1**).

**TABLE 1.** Cofactors examined for effect of Mask Mandates impact on COIVD-19 Mortality.

| DEMOGRAPHIC FACTORS |  |  | HEALTH RELATED FACTORS |  |
| --- | --- | --- | --- | --- |
| Population Factors | Social Capital Factors | Wealth & Housing Factors | Health Care Access Factors | Medical Comorbidities |
| Population total <sup>40</sup> | Social Capital Index (SCI); State-Level <sup>36</sup> | GDP State 2109 \$M <sup>41</sup> | Medicaid Expansion Yes/No <sup>42</sup> | State Health Score <sup>43</sup> |
| Children 0-18 <sup>51</sup> | SCI: Family Unity Subindex <sup>36</sup> | GDP \$/capita <sup>47</sup> | Insured by Employer <sup>44</sup> | Obesity <sup>45</sup> |
| Adults 19-25 <sup>51</sup> | SCI: Family Interaction Subindex <sup>36</sup> | Unemployment Rate % JAN 2020 <sup>46</sup> | Insured by Non-Group (Insured Selected) <sup>44</sup> | All Adults who reported Asthma <sup>47</sup> |
| Adults 26-34 <sup>51</sup> | SCI: Social Support Subindex <sup>36</sup> | Total Unemployment JAN 2020 <sup>45</sup> | Insured by Medicaid <sup>44</sup> | Men with Asthma <sup>42</sup> |
| Adults 35-54 <sup>51</sup> | SCI: Community Health Subindex <sup>36</sup> | Shelter Beds 2019 <sup>48</sup> | Insured by Military <sup>44</sup> | Women with Asthma <sup>42</sup> |
| Adults 55-64 <sup>51</sup> | SCI: Institutional Health Subindex <sup>36</sup> | Homeless 2019 <sup>49</sup> | Uninsured <sup>44</sup> | Adults with COPD/ Emphysema/ Bronchitis <sup>42</sup> |
| Adult 65+ <sup>51</sup> | SCI: Collective Efficacy <sup>36</sup> | Incarcerated per 100,000 adults <sup>50</sup> | Government Hospital beds/1000 <sup>51</sup> | Diabetes? Yes <sup>42</sup> |
| Ethnicity: White <sup>51</sup> | SCI: Philanthropic Health <sup>36</sup> | Resident Nursing Home per 100,000 adults <sup>52</sup> | Non-Profit Hospital beds /1000 <sup>46</sup> | Diabetes? Yes Pregnancy-Related <sup>42</sup> |
| Ethnicity: Black <sup>51</sup> | Social Capital Index; County-Level Method <sup>36</sup> | Density pop/sqmi <sup>53</sup> | For-Profit Hospital beds/1000 <sup>46</sup> | Diabetes? No <sup>42</sup> |
| Ethnicity: Hispanic <sup>51</sup> | SCI: County Population Weighted Index <sup>36</sup> | % Urban <sup>54</sup> | Total Hospital beds/1000 <sup>46</sup> | Diabetes? No Pre-Diabetes or Borderline Diabetes <sup>42</sup> |
| Ethnicity: Asian <sup>51</sup> |  | % Overcrowded Housing <sup>55</sup> | All Adults no doctor because of cost <sup>56</sup> | All Adults with Cardiovascular Disease <sup>42</sup> |
| Ethnicity: American Indian/Native Alaskan <sup>51</sup> |  | % Severely Overcrowded Housing <sup>40</sup> | Male no doc cost <sup>53</sup> | Men with Cardiovascular Disease <sup>42</sup> |
| Ethnicity: Native Hawaiian/Other Pacific Islander <sup>51</sup> |  |  | Female no doc cost <sup>53</sup> | Women with Cardiovascular Disease <sup>42</sup> |

Because Social Capital appears to mediate mortality effects independent of other cofactors ^34, 35^, an expanded set of Social Capital factors were included in the present study. The focus on state level outcomes in the United States meant that the Social Capital Index ^36^ was the metric of choice. These included both state level indexes, county level indexes, and county level population-weighted indexes. Additionally, for the state level Social Capital Index all five subindexes and two independent factors were included in the analysis (for more information, see the end of the Data subsection of the METHODS section).

The current study seeks to examine the impact of state-level Mask Mandates on the mortality of COVID-19 and then examine whether these 62 putative cofactors impacted the efficacy of that policy intervention.

There have been reports of political interference impacting the integrity of governmental scientific data including data on COVID-19 pandemic ^37,38^. These reports bring into questions the validity of that data. Because of this concern, in the current study, as in earlier studies ^4,15^, independently curated publicly available data was used ^39^.

## METHODS

### Hypotheses

The daily number of deaths in each state, from 30 days before to 50 days after the Mandate, was sourced from the New York Times COVID-19 Data Repository, due to concerns about the reliability of data collected by the federal government as noted above. The dates of the Mask Mandates’ implementation were obtained from the COVID-19 US State Policy (CUSP) Database ^57^. Mask Mandates for All Public Facing Employees were used, as in previous reports^4,15^.

Mask Mandates are hypothesized to reduce the number of COVID-19 deaths by reducing the number of people infected with the virus. People who die of COVID-19 in the immediate aftermath of a Mask Mandate being enacted would have already been infected with the virus before the Mandate took effect, due to the time-course of the infection and associated symptomology. A Mask Mandate is therefore expected to not immediately exert its maximum impact on the number of COVID-19 deaths: rather, a temporal lag (or “offset”) will be needed to assess the ultimate impact of the Mandate.

Analysis was conducted to identify the amount of time (number of days) required to attain full effectiveness in reducing the number of deaths due to COVID-19. Sensitivity analysis assessed the strength and stability of ODA models in training analysis and in leave-one-out (LOO) cross-generalizability analysis, sequentially removing post-Mandate data from 1 to 30 days after the Mandate was issued from the analysis. The date on which the maximum effect, measured by ESS, occurred for any given state was used as the lag period for that state.

After the efficacy of each state’s Mask Mandate was determined, the states whose Mandates had a statistically significant effect on deaths from COVID-19 were separated into categories according to the size of said effect. The categories were: relatively weak effect (ESS < 25%), a moderate effect (25% <= ESS < 50%), and a relatively strong effect (50% <= ESS < 75%), a strong effect (75% <= ESS < 90%), and a very strong effect (90% <= ESS).

An ODA was then performed to measure the influence of a variety of potential cofactors, using the ESS group of the states (the class variable), and a wide variety of potential cofactors (the attributes).

### Data

The daily number of new COVID-19 cases was obtained separately for each State in the 30 days before the Mandate was implemented, and in the 50 days after the Mandate was implemented. Case reports occurring before the Mandate were dummy-coded as class=0, and case reports occurring after the Mandate were coded as class=1. Data from The New York Times, based on reports from state and local health agencies, were initially downloaded from GitHub on January 8, 2021 (data re-confirmed on April 5, 2021) and cross referenced against the COVID-19 State Policy Database (updated March 3, 2021) for dates that Mandates were issued in each State.

Numerical attributes for the analysis of potential cofactors include Social Capital index (State-Level, Using County-Level Methods, and County-Population-Weighted Index, as well as various sub-indexes which are listed and briefly explained at the end of this subsection), pre-pandemic population, pre-pandemic GDP (per capita and total), age distribution (with brackets of 0-18, 18-25, 26-34, 35-54, 55-64, and 65+), pre-pandemic (January 2020) number of unemployed persons and unemployment rate, homelessness, shelter beds, incarceration number and rate, population density (people per square mile in 2015), urban overcrowding (number of houses having >1 person per room), severe urban overcrowding (number of houses having >1.5 people per room), percent population by ethnicity (White, Black, Hispanic, American Indian/Alaska Native, and Native Hawaiian/Other Pacific Islander categories), %population with obesity, health insurance status (divided into categories of Ensured by Employer, Ensured by Non-Group, Ensured by Medicaid, Ensured by Medicare, Ensured by Military, and Uninsured), number of hospital beds per thousand individuals (divided into categories of Government, Non-Profit Hospital, and For-Profit Hospital, as well as total beds per thousand), portion of adults who report not seeing a doctor due to cost (male, female, and all adults), adults who reported asthma (male, female, and all adults), adults told they Have COPD/Emphysema/Chronic Bronchitis, adults who reported having diabetes (with pregnancy related cases being counted separately, as well as those with borderline or pre-diabetes), and adults who reported cardiovascular disease (male, female, and all adults). The only categorical attribute included was if the state had expanded Medicaid (yes/no).

The subfactors of the Social Capital index and related indexes examined were: **Family Unity Subindex** (which includes percent of births in past year to unmarried women, percent of women 35-44 currently married (and not separated), and percent of children living in a single-parent family); **Family Interaction Subindex** (which includes percentage of parents reporting 4-plus hours per weekday of child TV time, percent of parents reporting 4-plus hours of child time on electronic devices, and percent of parents reporting someone read to child every day past week); **Social Support Subindex** (which includes percent saying get needed emotional support only (sometimes, rarely, or never), average number of close friends, percentage of neighbors doing favors for each other once-plus per month, and percent who trust all/most of neighbors); **Community Health Subindex** (which includes percent who volunteered with a group past year, percent who attended public meeting regarding community affairs, percent who attended meeting where political issues discussed, percent who worked with neighbors to improve/fix something, percent who served on a committee or as a group officer, percent who participated in demonstration, membership in an organizations per 1,000 residents, and non-profit organizations (including religious congregations) per 1,000 residents); **Institutional Health Subindex** (which includes, voting rate in the 2012 and 2016 presidential elections, mail-back response rate to 2010 census, percent with great confidence in corporations to do what’s right, percent with great confidence in the media, and percent with great confidence in public schools); the independent **Collective Efficacy Subindex** (which includes violent crimes per 100,000); and the independent **Philanthropic Health Subindex** (which includes the percent who made a donation of $25-plus dollars to a group in the past year).

## RESULTS

The analysis to determine the delay between the implementation of the Mandate and its effects revealed that the average delay was 22.8 days, with a standard deviation of 6.9 days. This is consistent with CDC estimates of 19 to 23 days between infection and death, depending on age^56^.

Hawaii had so few deaths that no analysis of its Mandate’s efficacy could be performed, and as such was not included in subsequent analyses. Of the 36 remaining states analyzed, 19 revealed a statistically significant MM effect, with one having a very strong effect (ESS LOO >= 90%), three having a strong effect (75% <= ESS < 90%), four having a relatively strong effect (50% <= ESS < 75%), eight having a moderate effect (25% <= ESS < 50%), and three having a weak effect (ESS < 25%) (SEE **TABLE 2**).

**TABLE 2.** A full half of States (19/37) showed a decrease in COVID-19 mortality with Mask Mandates. No States (0/37) showed an increase in COVID-19 mortality with Mask Mandates. Average Lag of 22.8 days with a standard deviation of 6.19 days was found in the 19 States with decreased mortality.

| State | Max ESS LOO | Max ESS LOO lag | Max ESS LOO group | Max ESS LOO P-value | ESS LOO for lag = 20 | ESS LOO P-value for lag = 20 |
| --- | --- | --- | --- | --- | --- | --- |
| MA | 92.32% | 27 | 4 | 1.11E-12 | 70.00% | 3.51E-08 |
| LA | 75.56% | 14 | 3 | 2.98E-10 | 73.33% | 5.69E-09 |
| MI | 76.36% | 28 | 3 | 2.35E-08 | 73.33% | 5.69E-09 |
| MN | 86.67% | 20 | 3 | 1.18E-12 | 86.67% | 1.18E-12 |
| IN | 53.81% | 29 | 2 | 0.000106 | 46.67% | 0.00024 |
| VT | 53.10% | 21 | 2 | 0.000013 | 50.00% | 0.000047 |
| NY | 63.33% | 27 | 2 | 4.59E-07 | 56.67% | 0.000001 |
| WA | 67.33% | 24 | 2 | 5.24E-07 | 62.64% | 0.000001 |
| ME | 38.57% | 29 | 1 | 0.006341 | 0.00% | 0.601727 |
| DE | 29.05% | 29 | 1 | 0.026932 | 20.00% | 0.085111 |
| NV | 48.33% | 30 | 1 | 0.000769 | 23.33% | 0.060227 |
| OR | 27.27% | 6 | 1 | 0.014631 | 23.33% | 0.05512 |
| FL | 30.00% | 20 | 1 | 0.017609 | 30.00% | 0.017609 |
| KY | 33.33% | 19 | 1 | 0.006349 | 32.76% | 0.007997 |
| WV | 45.00% | 30 | 1 | 0.00171 | 40.00% | 0.001665 |
| VA | 47.62% | 29 | 1 | 0.000872 | 43.33% | 0.000715 |
| AK | 17.93% | 13 | 0 | 0.037109 | 16.67% | 0.07275 |
| CA | 22.22% | 23 | 0 | 0.044729 | 20.00% | 0.062639 |
| TX | 23.53% | 16 | 0 | 0.004102 | 20.00% | 0.01186 |
| MS | 23.33% | 30 | NA | 0.065889 | 20.00% | 0.052082 |
| NM | 25.00% | 30 | NA | 0.069248 | 13.33% | 0.217408 |
| IL | 21.90% | 29 | NA | 0.104501 | -26.67% | 0.993651 |
| GA | 12.12% | 28 | NA | 0.183162 | -70.00% | 1 |
| OH | 15.33% | 25 | NA | 0.192575 | -26.67% | 0.997615 |
| NE | 10.00% | 30 | NA | 0.315334 | -73.33% | 1 |
| CO | 7.50% | 10 | NA | 0.346128 | -6.67% | 0.809173 |
| AZ | 8.33% | 30 | NA | 0.354973 | 3.33% | 0.5 |
| AR | 5.22% | 4 | NA | 0.385473 | 3.33% | 0.5 |
| PA | 3.33% | 26 | NA | 0.555556 | -3.33% | 0.881356 |
| DC | 0.39% | 16 | NA | 0.721726 | 0.00% | 0.754237 |
| NH | -11.67% | 30 | NA | 0.909677 | -16.67% | 0.99984 |
| UT | -4.67% | 0 | NA | 0.911865 | -10.00% | 0.973907 |
| NJ | -6.67% | 30 | NA | 0.941837 | -26.67% | 0.999601 |
| RI | -23.20% | 1 | NA | 0.999457 | -30.00% | 0.999475 |
| WY | 3.33% | * | NA | * | 0.00% | 0.754237 |
| MD | 3.33% | * | NA | * | 3.33% | 0.5 |
| HI | NA | NA | NA | NA | NA | NA |

The analysis of cofactors revealed that several cofactors were linked to an increased effect strength of the Mask Mandates in the 19 States in which Mandates had a statistically significant effect. Institutional Health Subindex of the Social Capital Index, State GDP, population density, State Health Score, portion of the population insured by non-group health insurance, and rate of pregnancy-related diabetes were the six factors which statistically significantly increased the efficacy of Mask Mandates (See **TABLE 3**). Conversely several cofactors were linked to a weaker effect strength of Mask Mandates. Factors that decreased the efficacy to the Mask Mandates were Incarceration rate, overcrowded housing (both regular and severe), portion of populations with health insurance from the military, portion of population without health insurance, portion of population who reported not seeing a doctor due to cost (regardless of gender) and, portion of the population who were ethnic American Indian/Native Alaskan were all statistically significantly linked to lower effect strength of Mask Mandates (SEE **TABLE 4**).

**TABLE 3.** A total of six factors increased the efficacy of Mask Mandates.

| **Factor INCREASED Efficacy of Mask Mandate** | **p-value** | **ESS** |
| --- | --- | --- |
| Institutional Health Subindex of SCI | 0.00836 | 59.38% |
| GDP State 2109 \$M | 0.04228 | 50.00% |
| Density ppl/sqmi | 0.0336 | 52.08% |
| Health Score | 0.02744 | 53.12% |
| Ensured by Non-Group | 0.02272 | 52.08% |
| Diabetes? Yes, Pregnancy-Related | 0.04692 | 42.71% |

**TABLE 4.** At total of six factors, plus three related cofactors, decreased the efficacy of Mask Mandates.

| **Factor DECREASED Efficacy of Mask Mandate** | **p-value** | **ESS** |
| --- | --- | --- |
| Incarcerated/100,000 adults | 0.02976 | 52.08% |
| % Over-Crowded Housing | 0.0308 | 51.04% |
| % Severely Over-Crowded Housing | 0.02904 | 51.04% |
| Insured by Military | 0.0038 | 59.38% |
| Uninsured | 0.02356 | 52.08% |
| All Adults no doctor because of cost | 0.03368 | 51.04% |
| Male no doctor cost | 0.03088 | 51.04% |
| Female no doctor cost | 0.03152 | 52.08% |
| Ethnicity: American Indian/Native Alaskan | 0.004664 | 37.50% |

## DISCUSSION

In over half of the States which implemented a Mask Mandate there was a statistically significant reduction in mortality due to COVID-19 infections. Moreover, in States that had no reduction in deaths, Mask Mandates did not increase COVID-19 mortality. Mask Mandates reduced death from COVID-19 in the majority and did no harm in any of the states studied. This shows mask mandates to be a highly effective governmental policy in the face of the COVID-19 pandemic.

The second result to be noted is the strikingly high degree of consistency between the empirically observed lag time demonstrated in this study and the clinically derived estimate of the average time lag between exposure and death from COVID-19. The independently derived estimate by the Centers for Disease Control and Prevention predicts a 19-to-23 day average duration of viral exposure to death in fatal cases of COVID-19 infection ^58^. The empirically determined lag between Mask Mandate and reduced mortality of 22.8 days is in complete agreement with the clinical guidance. The agreement between these lag times, determined by two different methods, nonetheless arriving at the same findings, bolsters the validity of the presented analysis.

Several factors had a statistically significant impact on the efficacy of mask mandates. The efficacy of mask mandates in reducing COVID-19 deaths was strengthened by a higher Social Capital Index’s Institutional Health Subindex, greater State GDP, greater Population Density, higher State Health Score, higher percentage with Non-Group Insurance, and higher rates of diagnosis of Gestational Diabetes. The Institutional Health Social Capital Subindex had the biggest positive impact on reduction in deaths. This important finding will be discussed further below.

As a generalization it appears as if greater wealth, health, and access to healthcare of choice improves the impact of Mask Mandates. At first glance the helpful influence of percent diagnosed with gestational diabetes seems paradoxical because diabetes is generally considered a risk factor for mortality with COVID-19. However, this finding may be an indicator of more proactive healthcare rather than of true increased rates of diabetes. Rates for gestational diabetes diagnoses vary by the screening method employed. Universal screening (which suggest a more proactive health system) compared with risk factor-based screening leads to more women being diagnosed with gestational diabetes ^59^. Rates of gestational diabetes may therefore be a proxy for broader health screening. Broader health screening reflects greater access to health care, which has been shown to improve MM efficacy. This inference is speculative but explains this otherwise paradoxical effect ^60^.

The factors which reduced the impact of mask mandates include dense living conditions (in jails or in overcrowded housing), military insurance, lack of insurance or lack of a doctor because of cost, as well as Native American/Native Alaskan ethnicity. Native Americans have the highest rate of poverty among any ethnic group ^61^. The link between poverty and worse health outcomes is well known ^62^. The presence of military insurance among factors decreasing the effect of mask mandates in reducing COVID-19 mortality could be a function of close living (like overcrowded housing), poverty, quality of health care, a military culture of denied vulnerability or some other factor.

Social capital improves the impact mask mandates. It is particularly interesting that specifically the Institutional Health Subindex is the dimension of social capital that impacts mask mandate effectiveness. One useful way of understanding of social capital is that it consists of three factors: bonding (relationships with friends and family), bridging (relationship between friends or families) and linking (relationship with a government official or agency) ^63^. Typically it is the more individual/ bonding factors of social capital like Family Unity and Family Interaction that impact health outcomes. The more societal/neighborhood aspects of social capital like Community Health and Institutional Health Subindexes are not particularly powerful in determining individual health ^64^. And yet, in our analysis, Institutional Health was a powerful factor in increasing the Mask Mandates’ efficacy in preventing deaths from COVID-19.

Institutional Health Subindex is one component of the Social Capital Index which is an indicator of confidence in institutions to do what is “right”. It is made up of a weighted average of votes in the presidential election per citizen (over 2012 and 2016), the Mail-back response rates for 2010 census and the “Confidence in Institutions Sub-Index” ^65^, a combination of subjects reporting at least some confidence in corporations, the media, and public schools. It is noteworthy that the impact of social capital Institutional Health index is so strong in this analysis. This finding reveals that trust in institutions which was especially, and perhaps unusually, important for saving lives in the COVID-19 pandemic in the United States.

Moreover, the negative impact of rancorous political debate might be more than merely increasing social discord. If the rancor erodes confidence in institutions, it may pose an actual threat to public health. In the context of the Covid-19 pandemic such political tactics may contribute to the loss of life.

As the process of vaccinating the US population unfolds, trust in public institutions plays an important role. Social Capital, especially general trust, affected vaccinations rates in the 2009 A(H1N1) flu outbreak in the United States ^66, 67^. Trust in public institutions (or lack thereof) is highly relevant to the public policies meant to limit the current COVID-19 pandemic as immunization is an essential requirement to get the populace to herd immunity.

## Data Availability

The New York Times (2020). Coronavirus
(Covid-19) data in the United States.

https://github.com/nytimes/covid-19-data

## AUTHOR NOTES

This study analyzed publicly available data and thus was exempt from Institutional Review Board review. No conflicts of interest were reported. The author would like to thank Dr. Alan Maloney for helpful edits and suggestions made in reviewing the manuscript.

## Notes

### Competing Interest Statement

The authors have declared no competing interest.

### Author Declarations

This study analyzed publically available data and thus was exempt from Institutional Review

## REFERENCES

1 Centers for Disease Control and Prevention, COVID-19 Response. COVID-19 Case Surveillance Public Data Access, Summary, and Limitations (version date: May 6, 2021). https://covid.cdc.gov/covid-data-tracker/#datatracker-home.

2 Kwok KO, Lai F, Wei WI, Wong SYS, Tang JWT. Herd immunity - estimating the level required to halt the COVID-19 epidemics in affected countries. J Infect. 2020;80(6):e32–e33. doi:10.1016/j.jinf.2020.03.027

3 Maloney MJ. The effect of face mask mandates during the COVID-19 pandemic on the rate of mask use in the United States. J Biochem Biotech 2020;S1:20–22.

4 Maloney MJ, Rhodes NJ and Yarnold, PR. Mask Mandates Can Rapidly and Efficiently Limit COVID-19 Spread: Month-Over-Month Effectiveness of Governmental Policies in Reducing the Number of New COVID-19 Cases in 37 US States and the District of Columbia. Optimal Data Analysis Vol. 9 (December 23, 2020), 237–254.

5 Coclite D, Napoletano A, Gianola S, Del Monaco A, D’Angelo D, Fauci A, Iacorossi L, Latina R, Torre G, Mastroianni CM, Renzi C, Castellini G, Iannone P. Face Mask Use in the Community for Reducing the Spread of COVID-19: A Systematic Review. Front Med (Lausanne). 2021 Jan 12;7:594269. doi: 10.3389/fmed.2020.594269

6 Joo H, Miller GF, Sunshine G, et al. Decline in COVID-19 Hospitalization Growth Rates Associated with Statewide Mask Mandates — 10 States, March–October 2020. MMWR Morb Mortal Wkly Rep 2021;70:212–216. DOI: http://dx.doi.org/10.15585/mmwr.mm7006e2

7 Erratum: Vol. 70, No. 6. MMWR Morb Mortal Wkly Rep 2021;70:293. DOI: http://dx.doi.org/10.15585/mmwr.mm7008a4

8 Van Dyke ME, Rogers TM, Pevzner E, et al. Trends in County-Level COVID-19 Incidence in Counties With and Without a Mask Mandate — Kansas, June 1–August 23, 2020. MMWR Morb Mortal Wkly Rep 2020;69:1777-1781. DOI: http://dx.doi.org/10.15585/mmwr.mm6947e2

9 Erratum: Vol. 69, No. 47. MMWR Morb Mortal Wkly Rep 2021;69:1663. DOI: http://dx.doi.org/10.15585/mmwr.mm695152a6

10 Sharoda Dasgupta, Ahmed M. Kassem, Gregory Sunshine, Tiebin Liu, Charles Rose, Gloria Kang, Rachel Silver, Brandy L. Peterson Maddox, Christina Watson, Mara Howard-Williams, Maxim Gakh, Russell McCord, Regen Weber, Kelly Fletcher, Trieste Musial, Michael A. Tynan, Rachel Hulkower, Amanda Moreland, Dawn Pepin, Lisa Landsman, Amanda Brown, Siobhan Gilchrist, Catherine Clodfelter, Michael Williams, Ryan Cramer, Alexa Limeres, Adebola Popoola, Sebnem Dugmeoglu, Julia Shelburne, Gi Jeong, Carol Y. Rao, Differences in rapid increases in county-level COVID-19 incidence by implementation of statewide closures and mask mandates — United States, June 1–September 30, 2020. Annals of Epidemiology 2021, ISSN 1047-2797, https://doi.org/10.1016/j.annepidem.2021.02.006

11 Timo Mitze, Reinhold Kosfeld, Johannes Rode, Klaus Wälde. Face masks considerably reduce COVID-19 cases in Germany. Proceedings of the National Academy of Sciences Dec 2020, 117 (51) 32293 32301; DOI: 10.1073/pnas.2015954117

12 Wei Lyu and George L. Wehby. Community Use Of Face Masks And COVID-19: Evidence From A Natural Experiment Of State Mandates In The US. Health Affairs 2020 39:8, 1419–1425

13 Joo H, Miller GF, Sunshine G, et al. Decline in COVID-19 Hospitalization Growth Rates Associated with Statewide Mask Mandates — 10 States, March–October 2020. MMWR Morb Mortal Wkly Rep 2021;70:212–216. DOI: http://dx.doi.org/10.15585/mmwr.mm7006e2

14 Erratum: Vol. 70, No. 6. MMWR Morb Mortal Wkly Rep 2021;70:293. DOI: http://dx.doi.org/10.15585/mmwr.mm7008a4

15 Maloney, MJ. Mask Mandate Prevented COVID-19 Deaths in Minnesota. Optimal Data Analysis Vol. 9 (October 26, 2020), 228–233.

16 Li L, Liu B, Liu SH, Ji J, Li Y. Evaluating the Impact of New York’s Executive Order on Face Mask Use on COVID-19 Cases and Mortality: a Comparative Interrupted Times Series Study. J Gen Intern Med. 2021 Jan 26:1–5. doi: 10.1007/s11606-020-06476-9

17 Albitar O, Ballouze R, Ooi JP, Sheikh Ghadzi SM. Risk factors for mortality among COVID-19 patients. Diabetes Res Clin Pract. 2020 Aug;166:108293. doi: 10.1016/j.diabres.2020.108293. Epub 2020 Jul 3. PMID: 32623035; PMCID: PMC7332436.

18 Atkins JL, Masoli JAH, Delgado J, Pilling LC, Kuo CL, Kuchel GA, Melzer D. Preexisting Comorbidities Predicting COVID-19 and Mortality in the UK Biobank Community Cohort. J Gerontol A Biol Sci Med Sci. 2020 Oct 15;75(11):2224–2230. doi: 10.1093/gerona/glaa183. PMID: 32687551; PMCID: PMC7454409.

19 Du RH, Liang LR, Yang CQ, Wang W, Cao TZ, Li M, Guo GY, D. J, Zheng CL, Zhu Q, Hu M, Li XY, Peng P, Shi HZ. Predictors of mortality for patients with COVID-19 pneumonia caused by SARS-CoV-2: a prospective cohort study. Eur Respir J. 2020 May 7;55(5):2000524. doi: 10.1183/13993003.00524-2020. Erratum in: Eur Respir J. 2020 Sep 24;56(3): PMID: 32269088; PMCID: PMC7144257.

20 “Predictors of mortality for patients with COVID-19 pneumonia caused by SARS-CoV-2: a prospective cohort study.” Rong-Hui Du, Li-Rong Liang, Cheng-Qing Yang, Wen Wang, Tan-Ze Cao, Ming Li, Guang-Yun Guo, Juan Du, Chun-Lan Zheng, Qi Zhu, Ming Hu, Xu-Yan Li, Peng Peng and Huan-Zhong Shi. Eur Respir J 2020; 55: 2000524. Eur Respir J. 2020 Sep 24;56(3):2050524. doi: 10.1183/13993003.50524-2020. Erratum for: Eur Respir J. 2020 May 7;55(5): PMID: 32973076.

21 Rajpal A, Rahimi L, Ismail-Beigi F. Factors leading to high morbidity and mortality of COVID-19 in patients with type 2 diabetes. J Diabetes. 2020 Dec;12(12):895–908. doi: 10.1111/1753-0407.13085. Epub 2020 Sep 2. PMID: 32671936; PMCID: PMC7405270.

22 Callender LA, Curran M, Bates SM, Mairesse M, Weigandt J, Betts CJ. The Impact of Pre-existing Comorbidities and Therapeutic Interventions on COVID-19. Front Immunol. 2020 Aug 11;11:1991. doi: 10.3389/fimmu.2020.01991. PMID: 32903476; PMCID: PMC7437504.

23 Chadeau-Hyam M, Bodinier B, Elliott J, Whitaker MD, Tzoulaki I, Vermeulen R, Kelly-Irving M, Delpierre C, Elliott P. Risk factors for positive and negative COVID-19 tests: a cautious and indepth analysis of UK biobank data. Int J Epidemiol. 2020 Oct 1;49(5):1454–1467. doi: 10.1093/ije/dyaa134. PMID: 32814959; PMCID: PMC7454561.

24 Burström, B., & Tao, W. (2020). Social determinants of health and inequalities in COVID-19. European journal of public health, 30(4), 617–618. https://doi.org/10.1093/eurpub/ckaa095

25 Price-Haywood EG, Burton J, Fort D, Seoane L. Hospitalization and Mortality among Black Patients and White Patients with Covid-19. N Engl J Med. 2020 Jun 25;382(26):2534–2543. doi: 10.1056/NEJMsa2011686. Epub 2020 May 27. PMID: 32459916; PMCID: PMC7269015.

26 Kabarriti R, Brodin NP, Maron MI, Guha C, Kalnicki S, Garg MK, Racine AD. Association of Race and Ethnicity With Comorbidities and Survival Among Patients With COVID-19 at an Urban Medical Center in New York. JAMA Netw Open. 2020 Sep 1;3(9):e2019795. doi: 10.1001/jamanetworkopen.2020.19795. PMID: 32975574; PMCID: PMC7519416.

27 Yehia BR, Winegar A, Fogel R, Fakih M, Ottenbacher A, Jesser C, Bufalino A, Huang RH, Cacchione J. Association of Race With Mortality Among Patients Hospitalized With Coronavirus Disease 2019 (COVID-19) at 92 US Hospitals. JAMA Netw Open. 2020 Aug 3;3(8):e2018039. doi: 10.1001/jamanetworkopen.2020.18039. PMID: 32809033; PMCID: PMC7435340.

28 Williamson EJ, Walker AJ, Bhaskaran K, Bacon S, Bates C, Morton CE, Curtis HJ, Mehrkar A, Evans D, Inglesby P, Cockburn J, McDonald HI, MacKenna B, Tomlinson L, Douglas IJ, Rentsch CT, Mathur R, Wong AYS, Grieve R, Harrison D, Forbes H, Schultze A, Croker R, Parry J, Hester F, Harper S, Perera R, Evans SJW, Smeeth L, Goldacre B. Factors associated with COVID-19-related death using OpenSAFELY. Nature. 2020 Aug;584(7821):430–436. doi: 10.1038/s41586-020-2521-4. Epub 2020 Jul 8. PMID: 32640463.

29 Elgar, F. J., Stefaniak, A., & Wohl, M. (2020). The trouble with trust: Time-series analysis of social capital, income inequality, and COVID-19 deaths in 84 countries. Social science & medicine (1982), 263, 113365. https://doi.org/10.1016/j.socscimed.2020.113365

30 Imbulana Arachchi J, Managi S. The role of social capital in COVID-19 deaths. BMC Public Health. 2021 Mar 3;21(1):434. doi: 10.1186/s12889-021-10475-8. PMID: 33657999; PMCID: PMC7928173.

31 Makridis, Christos and Wu, Cary, Ties that Bind (and Social Distance): How Social Capital Helps Communities Weather the COVID-19 Pandemic (May 4, 2020). Available at SSRN: https://ssrn.com/abstract=3592180 or http://dx.doi.org/10.2139/ssrn.3592180

32 Elgar FJ, Stefaniak A, Wohl MJA. The trouble with trust: Time-series analysis of social capital, income inequality, and COVID-19 deaths in 84 countries. Soc Sci Med. 2020 Oct;263:113365. doi: 10.1016/j.socscimed.2020.113365. Epub 2020 Sep 16. PMID: 32981770; PMCID: PMC7492158.

33 Makridis, Christos and Rothwell Jonathan T., The Real Cost of Political Polarization: Evidence from the COVID-19 Pandemic (June 29, 020). Available at SSRN: https://ssrn.com/abstract=3638373 or http://dx.doi.org/10.2139/ssrn.3638373

34 Giordano GN, Mewes J, Miething A. Trust and all-cause mortality: a multilevel study of US General Social Survey data (1978-2010). J Epidemiol Community Health. 2019 Jan;73(1):50–55. doi: 10.1136/jech-2018-211250. Epub 2018 Oct 15. PMID: 30322881; PMCID: PMC6839792.

35 Ye M, Aldrich DP. Substitute or complement? How social capital, age and socioeconomic status interacted to impact mortality in Japan’s 3/11 tsunami. SSM Popul Health. 2019 Apr 29;7:100403. doi: 10.1016/j.ssmph.2019.100403. PMID: 31080870; PMCID: PMC6506562.

36 U.S. Congress, Joint Economic Committee, Social Capital Project. “The Geography of Social Capital in America.” Report prepared by the Vice Chairman’s staff, 115th Cong., (2018). https://www.jec.senate.gov/public/index.cfm/republicans/2018/4/the-geography-of-social-capital-in-america#toc-005-backlink

37 Sharfstein JM. Science and the Trump Administration. JAMA. 2017;318(14):1312–1313. doi:10.1001/jama.2017.14813

38 Viglione G. “Four ways Trump has meddled in pandemic science - and why it matters”. Nature. 2020 Nov 3. doi: 10.1038/d41586-020-03035-4. Epub ahead of print. PMID: 33144734.

39 “Data from the New York Times, based on reports from state and local health agencies.” (2020). https://www.nytimes.com/interactive/2020/us/coronavirus-us-cases.html

40 The Kaiser Family Foundation State Health Facts. Data Source: Population, Age, Ethnicity (2019). https://www.kff.org/state-category/demographics-and-the-economy/population/

41 Bureau of Economic Activity (2019). Gross Domestic Product by State, 4th Quarter and Annual 2019 (PDF). www.bea.gov

42 The Kaiser Family Foundation State Health Facts. Data Source: Status of State Action on the Medicaid Expansion Decision (2020). https://www.kff.org/health-reform/state-indicator/state-activity-around-expanding-medicaid-under-the-affordable-care-act/?currentTimeframe=0&sortModel=%7B%22colId%22:%22Location%22,%22sort%22:%22asc%22%7D#note-4

43 America’s Health Rankings analysis of America’s Health Rankings composite measure, United Health Foundation (2018). AmericasHealthRankings.org. https://www.americashealthrankings.org/learn/reports/2018-annual-report/findings-state-rankings

44 The Kaiser Family Foundation State Health Facts (2019).Data Source: Health Insurance Coverage of the Total Population: Insured by Employer, Insured by Non-Group, Insured by Medicare, Insured by Medicaid, Insured by Military, Uninsured. https://www.kff.org/other/state-indicator/total-population/?currentTimeframe=0&sortModel=%7B%22colId%22:%22Location%22,%22sort%22:%22asc%22%7D

45 Centers for Disease Control and Prevention (CDC) (2018). CDC 2018 obesity map, Adult Obesity Prevalence Maps. https://www.cdc.gov/obesity/data/prevalence-maps.html

46 Bureau of Labor Statistics (2020). State Employment and Unemployment -- JANUARY 2020. https://www.bls.gov/news.release/archives/laus_03162020.htm

47 The Kaiser Family Foundation State Health Facts (2019). Data Source: Health Status: All Adults who reported Asthma, Men who reported Asthma, Women who reported Asthma, Share of Adults Ever Told They Have COPD/Emphysema/ Chronic Bronchitis, Diabetes? Yes, Diabetes? Yes Pregnancy-Related, Diabetes? No, Diabetes? No Pre-Diabetes or Borderline Diabetes, All Adults with Cardiovascular Disease, Men with Cardiovascular Disease, Women with Cardiovascular Disease. https://www.kff.org/state-category/health-status/

48 Department of Housing and Urban Development (2020). 2007 - 2019 HIC Data by State (XLSX). https://www.hudexchange.info/resource/3031/pit-and-hic-data-since-2007/

49 Department of Housing and Urban Development (2020). 2007 - 2019 PIT Counts by State (XLSX).https://www.hudexchange.info/resource/3031/pit-and-hic-data-since-2007

50 Kaeble D, Cowhig M and Bureau of Justice Statistics (2016). Correctional Populations In The United States, 2016. https://www.bjs.gov/index.cfm?ty=pbdetail

51 The Kaiser Family Foundation State Health Facts. Data Source: Government Hospital Beds, Non-Profit Hospital beds, For-Profit Hospital beds, Total Hospital beds (2019). https://www.kff.org/other/state-indicator/beds-by-ownership/?currentTimeframe=0&sortModel=%7B%22colId%22:%22Location%22,%22sort%22:%22asc%22%7D

52 Kaiser Family Foundation (2020). Total Number of Residents in Certified Nursing Facilities. https://www.kff.org/other/state-indicator/number-of-nursing-facility-residents/

53 United States Census Bureau (2010). Resident Population Data, Population Density. http://2010.census.gov/2010census/data/apportionment-dens-text.php

54 Iowa Community Indicators Program, Urban Percentage of the Population for States(2010). https://www.icip.iastate.edu/tables/population/urban-pct-states

55 Bennefield R, Bonnette R (2003). Structural and Occupancy Characteristics of Housing: 2000. Census 2000 Brief. https://www.census.gov/prod/2003pubs/c2kbr-32.pdf

56 The Kaiser Family Foundation State Health Facts. Data Source: Access to care: All Adults no doc cost; Male All Adult Males no doc cost All Adult Females no doc cost. (2019). https://www.kff.org/state-category/providers-service-use/access-to-care/

57 Raifman J, Nocka K, Jones D, Bor J, Lipson S, Jay J, and Chan P. (2020). “COVID-19 US state policy database.” Available at: www.tinyurl.com/statepolicies

58 Centers for Disease Control and Prevention, COVID-19 Pandemic Planning Scenarios (version date: September 10, 2020) https://www.cdc.gov/coronavirus/2019-ncov/hcp/planning-scenarios.html

59 Tieu J, McPhee AJ, Crowther CA, Middleton P, Shepherd E. Screening for gestational diabetes mellitus based on different risk profiles and settings for improving maternal and infant health. Cochrane Database Syst Rev. 2017 Aug 3;8(8):CD007222. doi: 10.1002/14651858.CD007222.pub4. PMID: 28771289; PMCID: PMC6483271.

60 Lagaert S, Snaphaan T, Vyncke V, Hardyns W, Pauwels LJR, Willems S. A Multilevel Perspective on the Health Effect of Social Capital: Evidence for the Relative Importance of Individual Social Capital over Neighborhood Social Capital. Int J Environ Res Public Health. 2021 Feb 5;18(4):1526. doi: 10.3390/ijerph18041526. PMID: 33562693; PMCID: PMC7914797.

61 United States Census Bureau (2016). American Community Survey. https://www.census.gov/programs-surveys/acs/

62 Chetty R, Stepner M, Abraham S, et al. The Association Between Income and Life Expectancy in the United States, 2001-2014. JAMA. 2016;315(16):1750–1766. doi:10.1001/jama.2016.4226

63 Kyne, D. and Aldrich, D.P. (2020), Capturing Bonding, Bridging, and Linking Social Capital through Publicly Available Data. Risk, Hazards & Crisis in Public Policy, 11: 61–86. https://doi.org/10.1002/rhc3.12183

64 Lagaert S, Snaphaan T, Vyncke V, Hardyns W, Pauwels LJR, Willems S. A Multilevel Perspective on the Health Effect of Social Capital: Evidence for the Relative Importance of Individual Social Capital over Neighborhood Social Capital. Int J Environ Res Public Health. 2021 Feb 5;18(4):1526. doi: 10.3390/ijerph18041526. PMID: 33562693; PMCID: PMC7914797.

65 Current Population Survey, September 2013: Volunteer Supplement conducted by the Bureau of the Census for the Bureau of Labor Statistics. Washington: Bureau of the Census. https://www2.census.gov/programs-surveys/cps/techdocs/cpssep13.pdf

66 Elgar FJ, Stefaniak A, Wohl MJA. The trouble with trust: Time-series analysis of social capital, income inequality, and COVID-19 deaths in 84 countries. Soc Sci Med. 2020 Oct;263:113365. doi: 10.1016/j.socscimed.2020.113365. Epub 2020 Sep 16. PMID: 32981770; PMCID: PMC7492158.

67 Rönnerstrand B. Contextual generalized trust and immunization against the 2009 A(H1N1) pandemic in the American states: A multilevel approach. SSM Popul Health. 2016 Sep 10;2:632–639. doi: 10.1016/j.ssmph.2016.08.004. PMID: 29349177; PMCID: PMC5757902.

